# Drivers of antibiotic use habits and animal disease incidence in smallholder livestock farms: evidence from a survey in Burkina Faso

**DOI:** 10.1101/2023.06.23.23291817

**Authors:** Eve Emes, Assèta Kagambèga, Michel Dione

## Abstract

Reducing nontherapeutic antibiotic (ABU) use in livestock animals has been identified as an important way of curbing the growth of antimicrobial resistance (AMR). However, this ABU can be an important disease management tool, and farmers may not feel safe to reduce it without simultaneous interventions to safeguard animal health. It is therefore important to determine a) if nontherapeutic ABU is important for averting livestock animal disease, b) which factors can encourage farmers to improve antibiotic stewardship on their own terms, and c) which factors can be paired with ABU reduction in order to safeguard against any animal health risks.

We investigated these questions using data from the AMUSE survey, which is designed to evaluate knowledge, attitudes and practices relating to AMR in smallholder livestock farms. Our sample covered 320 animal herds from 216 smallholder livestock farms in Burkina Faso, with species including poultry, small ruminants, and cattle. The determinants of the likelihood of animal disease and nontherapeutic ABU were investigated using logistic regression.

We found that nontherapeutic ABU was positively associated with animal disease, although the potential endogeneity of this relationship should be investigated further. We also found that going primarily to a public veterinarian for animal health services was associated with a lower likelihood of nontherapeutic ABU. We also found some evidence that going to public veterinarians, and a higher level of formal education, were associated with a lower likelihood of animal disease.

These findings support the expansion of public veterinary services as a way to encourage antibiotic stewardship, and to safeguard against any animal health risks associated with ABU reduction.

## Introduction

Antimicrobial resistance (AMR), the ability of microbial pathogens to survive in the presence of antimicrobials, is an important and growing danger to human health, environmental health, and food security. The use of antimicrobials (AMU) by humans has resulted in growing rates of AMR(1). The use of antibiotics in livestock animals is one of the biggest forms of AMU, and has been the target of a lot of national and international health policy initiatives(2,3). In particular, international AMR policy targets a reduction in ‘irrational’ AMU in livestock animals, usually referring to nontherapeutic (metaphylactic, prophylactic and growth-promoting) use(4–6).

However, characterising these uses as irrational is neither fair nor constructive. There is good evidence of health benefit from sub-inhibitory doses of antibiotics(7), and our previous work has pointed to nontherapeutic antibiotic use averting animal disease in smallholder livestock farms(8). In addition to this, the potential growth-promoting effects of antibiotic use in livestock animals may be important for smallholder farmers’ incomes, and for general food security. This is especially important for countries such as Burkina Faso, which has both a high rate of population growth and a relatively low degree of food security(9,10). In addition to this, smallholder livestock farmers exist as part of a network of economic interdependencies which involves marketeers, suppliers, creditors, landlords, pharmaceutical sellers, animal health professionals, and others(11). Simply placing legal restrictions on the use of antibiotics in these farms may not be feasible, and could result in farmers circumventing restrictions by buying substandard or counterfeit antibiotics illegally, which may worsen AMR outcomes.

For these reasons, it is important to determine three main things. Firstly, the extent to which nontherapeutic antibiotic use in smallholder livestock farms is important for averting animal disease. Secondly, which non-antibiotic measures can avert animal disease, which could be combined with antibiotic use reduction to mitigate risks. And finally, which other factors can encourage farmers to use fewer nontherapeutic antibiotics on their own terms.

In order to address these questions, we analysed data collected using the AMUSE survey among livestock farmers who preliminarily kept chicken in peri-urban areas of Ouagadougou. AMUSE is a standardised survey developed by the International Livestock Research Institute to assess knowledge, attitudes and practices (KAP) relating to antibiotic use and resistance in smallholder livestock farms(12). The survey has been used in Burkina Faso(13), Ethiopia(14), Senegal(15,16), and Uganda(8,17), and adds to a growing bank of knowledge which can inform agricultural AMR policies at the national and international level. The survey allows results to be compared across contexts, and we have used these survey data to write papers similar to this one focusing on Senegal(15) and Uganda(8).

The poultry sector plays a major role in the socio-economic development of Burkina Faso. It is among the country’s most dynamic sectors. The value of poultry production in 2011 was estimated at over XOF 85 billion (USD 140 million), representing about 6% of the country’s agricultural Gross Domestic Product (GDP). In 2016, intensive farm numbers were estimated at 868,450 layers and 70,605 broilers (including cocklepers)17. Modern livestock farming is mainly concentrated around certain large cities such as Ouagadougou(18). The size of the flocks varies from 140 to more than 1000 birds, with more farmers focusing on meat production than eggs(13). However most farmers also keep other livestock such as cattle, small ruminants (sheep and goats).

## Ethical approval

The study was approved by the ethical committee of the Ministry of Health, Burkina Faso, with reference number 2020-9-186. Informed (written and signed) consent was obtained from each participant before they were interviewed. Consequently, all participants gave their consent to participate in the study.

## Methods

### Survey methods

We used secondary data from a survey implemented in Burkina Faso between March and July 2020 that aims to evaluate knowledge, attitudes and practices of poultry farmers on the use of veterinary drugs with a focus on antibiotics in urban and peri-urban poultry farmers in Ouagadougou, Burkina Faso(13). During and after data collection, authors had access to information (including name and gender) which could identify individual participants.

### Study area

Ouagadougou is the most densely populated city in Burkina Faso, West Africa, with 2,415,266 inhabitants, and considering the high demand for meat and eggs, poultry farmers settle in and around the city to meet this need. A total of 216 poultry farms (broilers and layers) were selected for the study. From each farm, the manager (the owner or designated worker) was requested to participate in the study.

**Figure 1.**
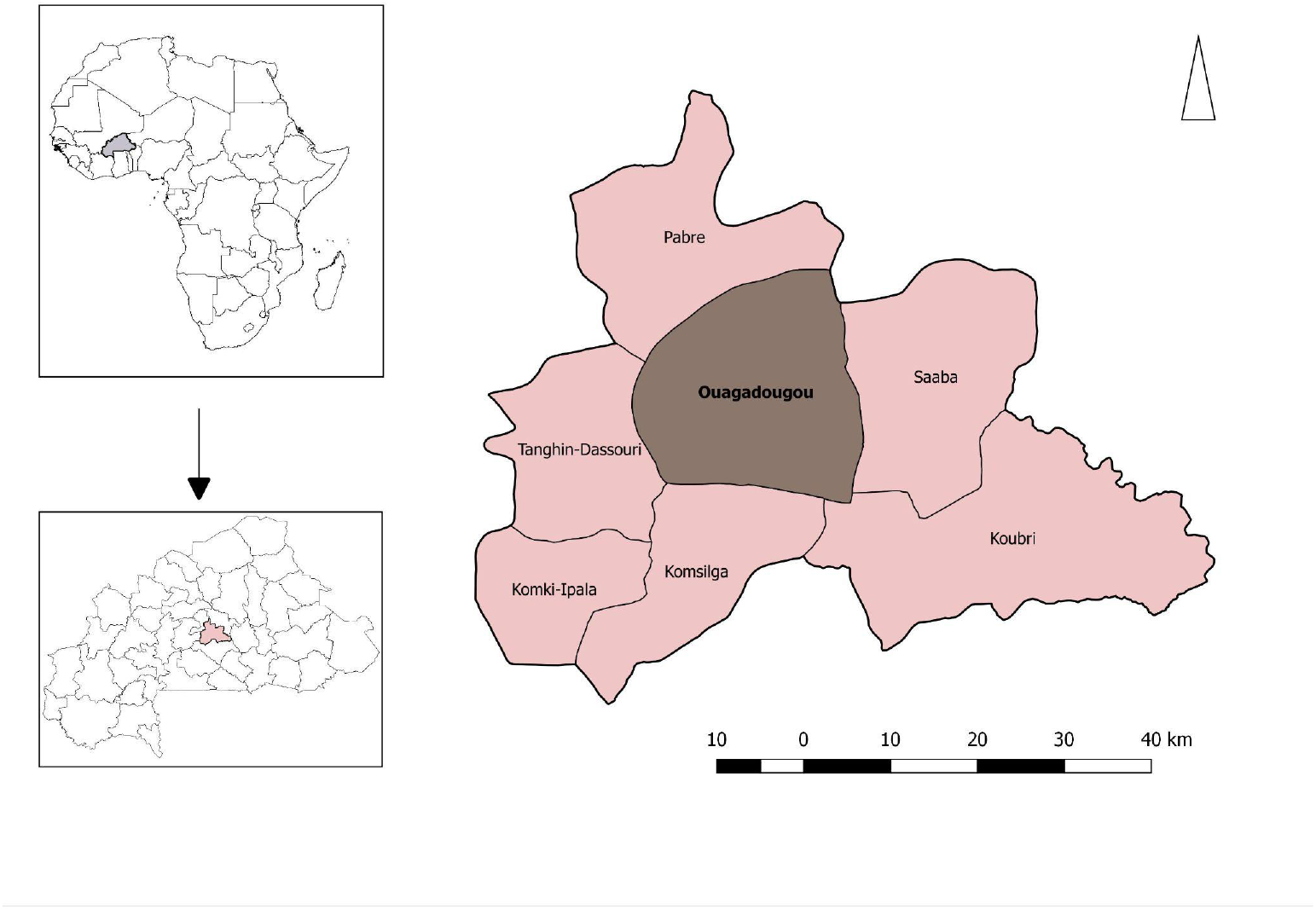
map of study area

### Sample size calculation

The sample size was calculated using the following conditions: in a study in Ghana (of similar production systems), expected proportion of 12.1% of farmers who adopted good practices (farmers sought individual prescriptions from the veterinary office before purchasing drugs for the birds)(19), a risk of error α of 5%, and a confidence level of 95%. Based on this estimate, the required sample size was 163 poultry farms. This number was increased to 216 to account for missing data and the size of the previous study.

### Statistical methods

## Overview

We compiled the survey responses from each farm into a cross-sectional dataset. Where farms had multiple flocks and herds of different species, we treated each flock or herd as a separate unit of analysis, given that much of the information collected was species-specific.

## Difficulties with the dataset

We encountered a number of difficulties while cleaning and preparing the data. Initially, we had planned to look at three outcomes: the occurrence of disease in the flock or herd in the last six months, the use of antibiotics for nontherapeutic purposes, and the use of antibiotics intended for humans in animals. However, only one farm reported using human antibiotics in animals, so we did not use this as an outcome variable.

We also intended to investigate the effect of frequency of use of vaccinations, antibiotics and other drugs on the occurrence of disease. However, data on the use of these drugs only covered the last four weeks, meaning that there would be considerable endogeneity between use of these drugs and the occurrence of disease (if the drugs were used in response to incidents of disease) and may not be reflective of general use habits. However, we did have data on whether or not farmers habitually used antibiotics for nontherapeutic purposes, and on their beliefs about the purposes for which antibiotics can be used.

We had intended to look at the effect of farmers having taken part in campaigns (e.g. public vaccination campaigns, or information campaigns run by NGOs) on antibiotic use and disease. However, only 7 farms reported having taken part in such campaigns in the last year, so we were unable to use this as a covariate.

Finally, we originally wanted to look at all of the livestock species represented in the sample (chicken and other poultry, pigs, cows, rabbits, sheep and goats, and horses and donkeys). However, several species were present only on a small number of farms, and so we restricted our analysis to chickens, cows, and goats and sheep.

## Specifications

After cleaning and exploring the data, we arrived at two outcomes of interest: the occurrence of disease in the flock or herd in the last six months, and the nontherapeutic use of antibiotics (defined here as use for prophylaxis and fattening). Note that the subset of farms which used antibiotics prophylactically was identical to the subset that used antibiotics nontherapeutically, so we will use these two terms interchangeably.

We used logistic regression to investigate the effect of our covariates on the likelihood of our outcomes. In all specifications, we controlled for the number of animals in the flock or herd. We displayed our results separated by species, and then for the sample as a whole.

We first looked at the effect of antibiotic use habits and attitudes on the likelihood of disease. Our covariates were: habitually using antibiotics prophylactically, and believing that antibiotics can be used for fattening.

We then looked at the effect of the level of formal education and the main provider of animal health services on the same outcome. For our first covariate, we assigned each farmer a score based on their level of formal education (0: no education, 1: literacy only, 2: primary education, 3: secondary education, 4: university education). A small number of farmers reported having undergone vocational or informal training, but because these forms of training could not be directly compared with formal education we did not assign them a score. The categories of animal health service provider looked at were: community animal health worker, public veterinarian, qualified private veterinarian, and private veterinarian with no or unknown qualifications. Farmers reported going to other providers (e.g. traditional healers, friends and neighbours), but too few farms reported using them for them to be included as covariates.

Thirdly, we looked at the determinants of habitually using antibiotics for nontherapeutic purposes. Our covariates here were: the level of formal education of the farmer, the main provider of animal health services, and whether or not the farmer usually went to a professional (from any of the aforementioned categories) for diagnosis and treatment.

After running these main results, we performed additional robustness checks. In our main results, we had found that both public vet access and nontherapeutic AMU seemed to influence the likelihood of disease, and that public vet access also seemed to influence the likelihood of nontherapeutic AMU. To investigate whether or not public vet access influenced the likelihood of disease independently of any effect on the likelihood on nontherapeutic AMU, we regressed this outcome against both covariates together.

As an additional robustness check, we redid our main results (and the above specification) for the whole sample while controlling for the farm species. We did this to ensure that any results which emerged from these specifications (looking at the whole sample) was not simply the result of differences in outcomes between farms of different species. For example, if chicken farms used the most prophylactic antibiotics and also naturally had the highest incidence of disease, it may falsely appear that prophylactic AMU contributes to disease.

## Results

### Main results

**Table 1.** effect of antibiotic use habits and attitudes on the likelihood of disease (odds ratio)

|  | Likelihood of Disease in Last 6 Months |  |  |  |
| --- | --- | --- | --- | --- |
|  | Chickens and other Poultry<br>(1) | Goats and Sheep<br>(2) | Cattle<br>(3) | Whole Sample<br>(4) |
| uses antibiotics prophylactically | 0.599<br>t = -0.786 | 20.654<br>t = 3.260*** | 4.043<br>t = 1.767* | 5.454<br>t = 5.791*** |
| believes that antibiotics can be used for fattening | 1.404<br>t = 0.839 | 0.645<br>t = -0.455 | 1.054<br>t = 0.073 | 0.975<br>t = -0.090 |
| number of animals in flock / herd | 1.000<br>t = -0.029 | 1.008<br>t = 0.378 | 1.098<br>t = 2.017** | 1.001<br>t = 3.253*** |
| Constant | 8.213<br>t = 3.307*** | 0.069<br>t = -2.810*** | 0.082<br>t = -3.039*** | 0.428<br>t = -3.362*** |
| N | 212 | 59 | 49 | 320 |
| Log Likelihood | -83.991 | -19.891 | -25.624 | -164.051 |
| Akaike Inf. Crit. | 175.982 | 47.782 | 59.248 | 336.102 |

Notes:
\*\*\* Significant at the 1 percent level.
\*\* Significant at the 5 percent level.
\* Significant at the 10 percent level.

In this specification, habitual prophylactic use of antibiotics was associated with a greater likelihood of disease in the last 6 months for goat, sheep and cattle farms, and for the sample as a whole, but not for poultry farms.

**Table 2.** effect of formal education and main animal health service provider on the likelihood of disease (odds ratio)

|  | Likelihood of Disease in Last 6 Months |  |  |  |
| --- | --- | --- | --- | --- |
|  | Chickens and other Poultry<br>(1) | Goats and Sheep<br>(2) | Cattle<br>(3) | Whole Sample<br>(4) |
| formal education level score | 0.836<br>t = -0.988 | 0.739<br>t = -0.931 | 0.696<br>t = -1.224 | 0.875<br>t = -1.210 |
| goes primarily to community animal health worker | 1.734<br>t = 0.833 | 0.481<br>t = -0.701 | 0.388<br>t = -0.825 | 0.944<br>t = -0.135 |
| goes primarily to public vet | 0.712<br>t = -0.509 | 0.175<br>t = -1.267 | 0.104<br>t = -1.696* | 0.328<br>t = -2.414** |
| goes primarily to unqualified private vet | 1.512<br>t = 0.446 | 0.712<br>t = -0.280 | 0.125<br>t = -1.331 | 0.674<br>t = -0.752 |
| goes primarily to qualified private vet | 1.385<br>t = 0.384 | 0.556<br>t = -0.404 | 0.263<br>t = -0.857 | 0.569<br>t = -1.019 |
| number of animals in flock / herd | 1.000<br>t = 0.019 | 1.008<br>t = 0.298 | 1.129<br>t = 2.296** | 1.002<br>t = 5.694*** |
| Constant | 7.867<br>t = 3.077*** | 0.577<br>t = -0.477 | 0.486<br>t = -0.651 | 1.488<br>t = 0.920 |
| N | 206 | 58 | 48 | 312 |
| Log Likelihood | -77.857 | -24.816 | -22.672 | -170.991 |
| Akaike Inf. Crit. | 169.714 | 63.631 | 59.344 | 355.982 |

Notes:
\*\*\* Significant at the 1 percent level.
\*\* Significant at the 5 percent level.
\* Significant at the 10 percent level.

We did not find any association between the level of formal education and the likelihood of disease in the last six months in any specification. Going primarily to a public vet was associated with a lower likelihood of disease for cattle farms and for the sample as a whole, but going primarily to any of the other animal health service provider types was not significantly associated with the likelihood of disease.

**Table 3.** determinants of habitually using antibiotics for nontherapeutic purposes (odds ratio)

|  | Likelihood of Using Antibiotics Nontherapeutically |  |  |  |
| --- | --- | --- | --- | --- |
|  | Chickens and other Poultry<br>(1) | Goats and Sheep<br>(2) | Cattle<br>(3) | Whole Sample<br>(4) |
| formal education level score | 1.176<br>t = 0.919 | 1.046<br>t = 0.135 | 1.006<br>t = 0.019 | 1.174<br>t = 1.336 |
| goes primarily to community animal health worker | 8.598<br>t = 2.101** | 0.502<br>t = -0.521 | 1.372<br>t = 0.238 | 1.063<br>t = 0.131 |
| goes primarily to public vet | 0.493<br>t = -0.757 | 1.529<br>t = 0.319 | 0.445<br>t = -0.547 | 0.184<br>t = -3.162*** |
| goes primarily to unqualified private vet | 1.211<br>t = 0.190 | 0.632<br>t = -0.302 | 0.585<br>t = -0.338 | 0.475<br>t = -1.311 |
| goes primarily to qualified private vet | 1.787<br>t = 0.541 | 0.375<br>t = -0.541 | 0.531<br>t = -0.366 | 0.393<br>t = -1.507 |
| professional provides diagnosis and treatment | 0.274<br>t = -1.805* | 5.761<br>t = 2.291** | 2.906<br>t = 1.253 | 1.737<br>t = 1.706* |
| number of animals in flock / herd | 1.001<br>t = 2.227** | 0.977<br>t = -0.792 | 0.969<br>t = -0.554 | 1.002<br>t = 6.221*** |
| Constant | 3.683<br>t = 1.795* | 0.314<br>t = -0.887 | 0.311<br>t = -0.912 | 0.610<br>t = -1.101 |
| N | 206 | 58 | 48 | 312 |
| Log Likelihood | -66.265 | -26.935 | -22.862 | -149.550 |
| Akaike Inf. Crit. | 148.530 | 69.870 | 61.724 | 315.101 |

Notes:
\*\*\* Significant at the 1 percent level.
\*\* Significant at the 5 percent level.
\* Significant at the 10 percent level.

Here, formal education level did not seem to be significantly related to nontherapeutic antibiotic use. From among the main providers of animal health services, going primarily to a community animal health worker was associated with a higher likelihood of nontherapeutic antibiotic use (for poultry farms) whereas going primarily to a public vet was associated with a lower likelihood (for the sample as a whole). Having an animal health professional (of any kind) provide diagnosis and treatment was associated with a lower likelihood of nontherapeutic antibiotic use for poultry farms, and a higher likelihood for goat and sheep farms and for the sample as a whole.

### Robustness

**Table 4:**
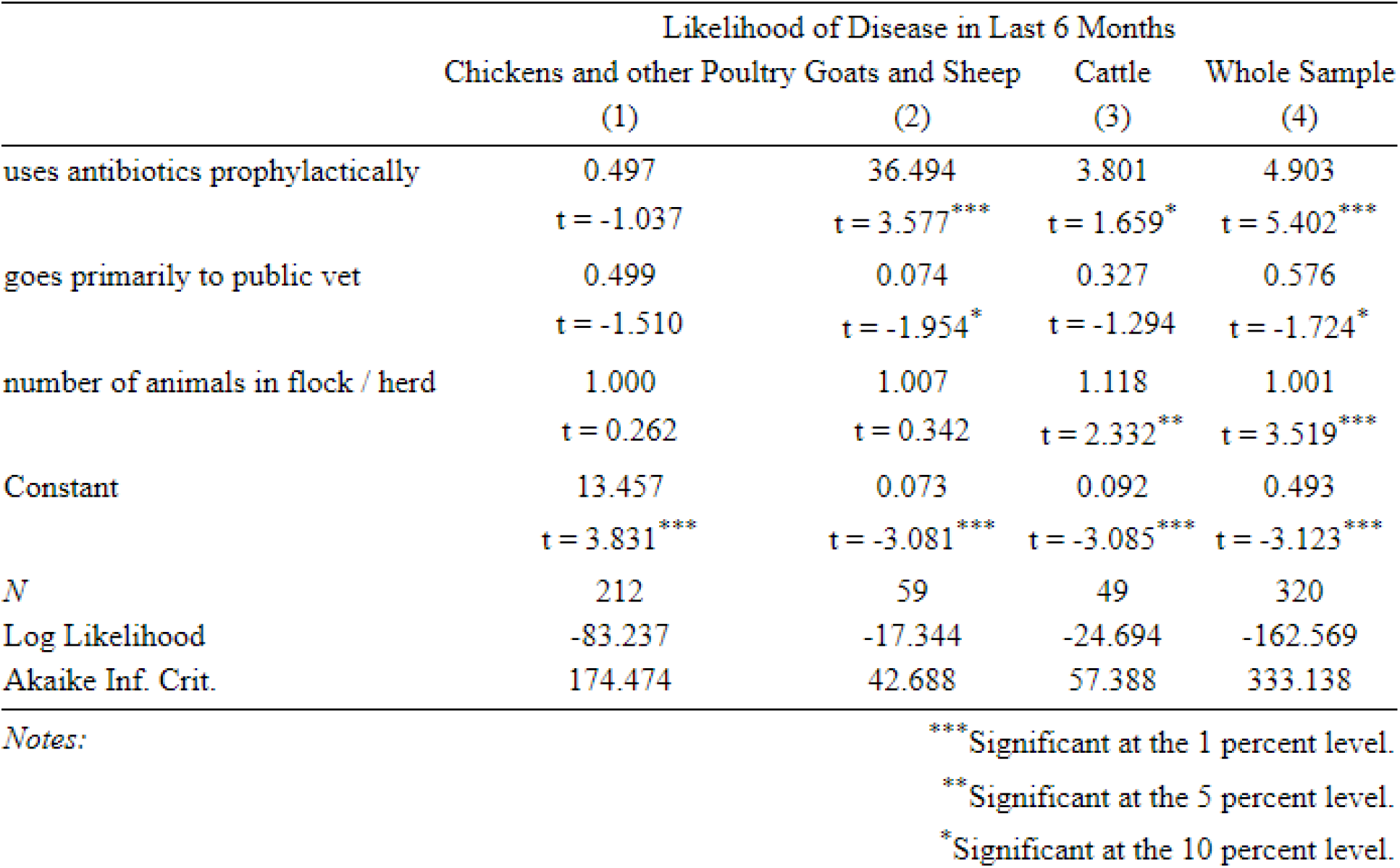
separating the effect of prophylactic AMU and public vet access on disease incidence (odds ratio)

In our main results, we found that going primarily to a public vet was associated with a lower likelihood of disease (for cattle farms and for the sample as a whole). Public vets were also associated with a lower likelihood of using antibiotics prophylactically (for the sample as a whole), a practice which in turn was positively associated with the likelihood of disease. We can therefore not be sure if public vets lower the likelihood of disease solely via their effect on prophylactic antibiotic use, or if they have an independent effect on disease incidence. For this reason, we regressed the likelihood of disease against both covariates together.

We found that the expected relationship remained significant in the case of both covariates. It appears that going to a public vet is associated with a lower likelihood of disease independently of its effect on prophylactic antibiotic use.

**Table 5:**
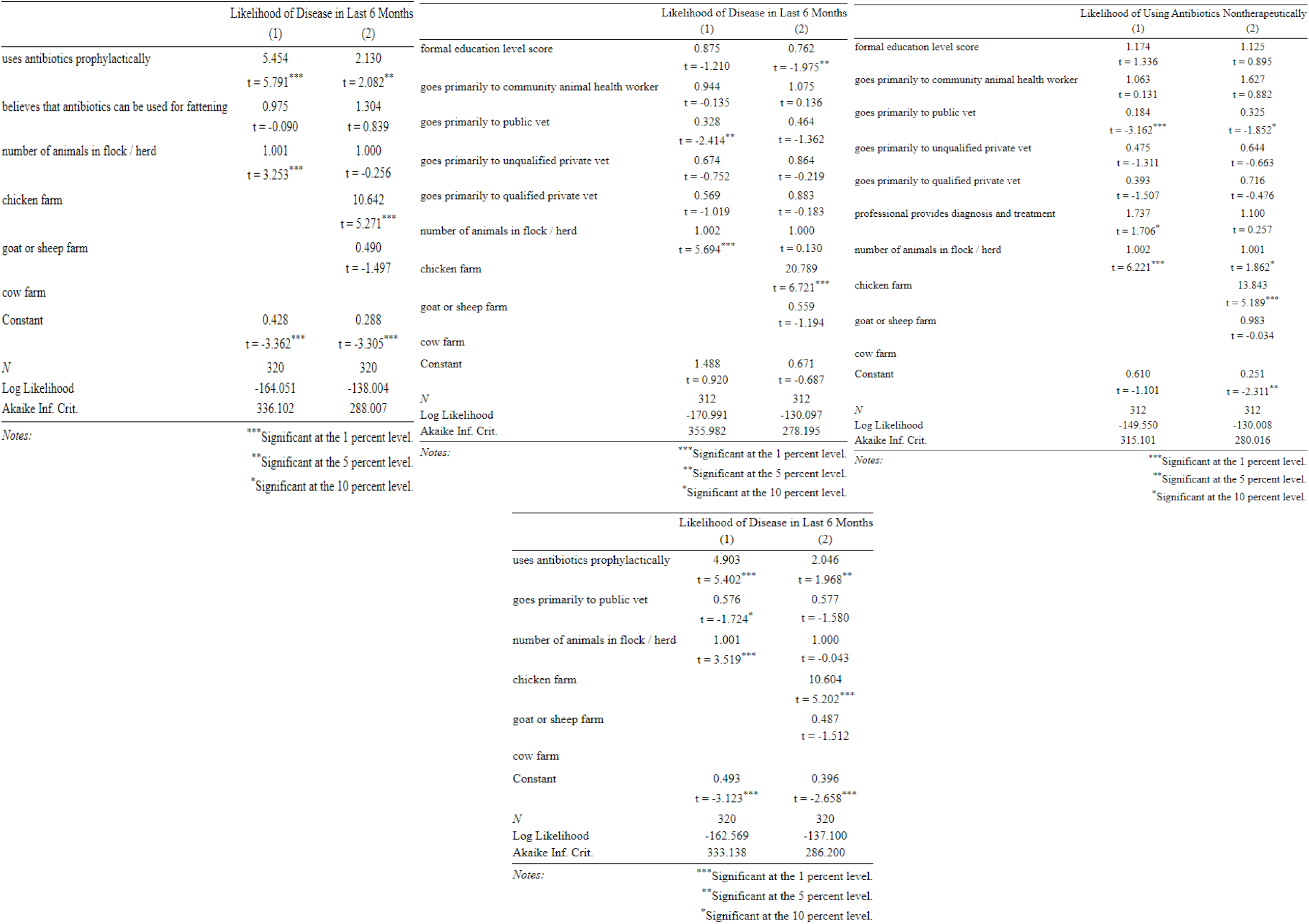
redoing main specifications while controlling for farm species (odds ratio)

When controlling for the animal species of each flock / herd, prophylactic antibiotic use remained positively associated with the likelihood of disease. However, the size of the effect was smaller than in the original specification, with some of it likely being mediated by the simultaneously higher likelihood of disease and higher prophylactic antibiotic use found on chicken farms.

When controlling for the flock / herd species, formal education score now seemed to be significantly associated with a lower likelihood of disease, and going to a public vet was no longer associated with a lower likelihood of disease. The chicken dummy was the only species dummy significantly related to the outcome variable, and chicken farms were no more or less likely to go to a public vet, so this loss of significance is likely to have arisen from overspecification rather than from endogeneity with flock / herd species.

With controls for farm species, going to a public vet continued to be associated with a lower likelihood of nontherapeutic antibiotic use, and professionals providing diagnosis and treatment was no longer associated with a higher likelihood of nontherapeutic antibiotic use.

Finally, when looking at the effect of public vets and nontherapeutic antibiotic use on likelihood of disease, the addition of species dummies reduced the size of the relationship between prophylactic antibiotic use and likelihood of disease, but did not change the sign or negate its statistical significance. Going to a public vet was no longer significantly associated with a lower likelihood of disease, however the coefficient did not change, suggesting that this change was due to a loss of statistical power rather than to endogeneity in the original specification.

## Discussion

### Results and interpretation

The two consistent and robust trends identified from our results were a) that habitual prophylactic antibiotic use was associated with a higher likelihood of animal disease, and b) that primarily going to a public veterinarian for animal health services was associated with a lower likelihood of nontherapeutic antibiotic use. In addition to these findings, we found more sporadic evidence to suggest that going to a public veterinarian and having a higher level of formal education were associated with a lower likelihood of animal disease.

### Determinants of likelihood of disease

It is interesting that habitual prophylactic antibiotic use was positively associated with animal disease here. There could be endogeneity in the sense that having had more animal disease in the last 6 months may have prompted farmers to adopt more cautious antibiotic use protocols which involve greater prophylactic use. This result contradicts what we have found in similar studies using the AMUSE survey tool: in Uganda(8) we found that prophylactic AMU guarded against disease, and in Senegal(15) we found that higher antibiotic use was associated with stronger broiler productivity, suggesting some growth-promotion or sub-inhibitory health benefit. This result would have to be verified using farm-level trials of antibiotic use reduction before being able to draw the conclusion that reducing AMU on farms can be done without putting animal health at risk.

We also found more sporadic evidence that primarily accessing public veterinarians may be associated with a lower likelihood of disease. While this echoes our findings from Uganda that access to animal health services consistently improved disease outcomes(8), it is interesting to note that not all providers of animal health services were seen to improve disease outcomes. This suggests that future interventions to guard against animal health risks while reducing on-farm AMU should prioritise access to, and funding for, public vets.

The fact that private vets, regardless of qualification status, did not seem to improve disease outcomes raises questions about the potential for perverse incentives in private antibiotic prescribing. For example, there may be an incentive to sell inappropriate but expensive medicines.

There was also some suggestion that a higher level of formal education reduced the likelihood of disease, but the mechanism of causality would have to be explored by people with knowledge of the public education system before conclusions can be drawn.

## Determinants of likelihood of nontherapeutic antibiotic use

We found robust evidence that going primarily to a public vet for animal health services was associated with a lower likelihood of using antibiotics nontherapeutically. The fact that there was some evidence of accessing these services being associated with a lower disease incidence as well could be indicative of a few things. For one, if the former relationship is causal, then any disease-averting effects of public vets may operate through a reduction in nontherapeutic AMU, which we found to be positively associated with disease.

We tested this idea, regressing the likelihood of disease against prophylactic antibiotic use and public vet access together. We found that public vet access continued to be associated with disease incidence in the same way, with the effect size falling somewhat. When controlling for flock / herd species, the relationship with prophylactic antibiotic use remained intact, while the relationship with public vet access narrowly lost statistical significance.

However, the coefficient remained unchanged with the added controls, suggesting that this might have been due to overspecification. This suggests that public vet access may reduce the likelihood of animal disease, both inherently and via encouraging a reduction in nontherapeutic antibiotic use.

It is also interesting to note that going to community animal health workers seemed to be associated with a greater likelihood of nontherapeutic antibiotic use in chickens, and that going to professionals (of any kind) for diagnosis and treatment had an uncertain effect on the likelihood of nontherapeutic AMU. This could suggest that animal health professionals do not, by default, prioritise antibiotic stewardship. This is of course quite fair -faced with the immediacy of curing livestock animals and averting disease, they may choose to prioritise animal health by encouraging greater antibiotic use. The fact that public vets did not follow this trend could mean that they have been more exposed to government goals as part of the ongoing NAP: these include a drive to involve veterinary medicine in antibiotic stewardship efforts and to change prescribing culture(20). In the case of private vets especially, there may also be an incentive to overprescribe to maximise revenue, or to prescribe excessively broad-spectrum antibiotics to minimise the risk of ineffective treatment which may harm future business.

### Implications for research and practice

This study, and similar studies conducted using the AMUSE survey tool, have aimed to inform interventions which can facilitate reductions in on-farm antibiotic use while safeguarding against any animal health risks associated with doing so. Our findings support the expansion of public veterinary services as a potential way of achieving both of these outcomes in Burkina Faso. Regarding the relationship observed between animal disease and nontherapeutic antibiotic use, implications for practice are difficult to draw because of the potential for endogeneity. Further investigation can give us a better understanding of the advisability and safety of reducing on-farm AMU. The sporadic evidence that formal education may improve disease outcomes also merits further exploration.

These results cannot in themselves tell us which interventions to implement, but they do provide a useful guideline on future avenues to explore. Given our findings here, future farm-level trials could investigate the mechanisms by which public veterinary services encourage better stewardship, and if (and how) they guard against animal disease. These trials could lay the groundwork for future improvements to public veterinary services, and for interventions which combine antibiotic use reduction with the expansion of access to these services. Future mixed-methods research could also be done to investigate the ways in which education interacts with farm practices and disease outcomes, and also to look into how the incentives faced by private prescribers may interact with stewardship and animal health concerns.

In any research concerning antibiotic stewardship, we must also remember that smallholder farmers exist as part of a complex network of actors which includes lenders, landlords, drug sellers, animal health professionals, marketeers and more(11). Any intervention aiming to improve stewardship outcomes must acknowledge and involve this entire network.

## Conclusions

Using a survey of smallholder livestock farms in Burkina Faso, we found that there was a greater likelihood of animal disease in farms which habitually used antibiotics for nontherapeutic purposes, although this relationship may be subject to endogeneity. We also found that going primarily to a public veterinarian for animal health services (as opposed to other animal health professionals) was associated with a lower likelihood of using antibiotics nontherapeutically. We also found some evidence that a higher level of formal education, and primarily going to public veterinarians, was associated with a lower likelihood of animal disease.

These findings support the expansion of public veterinary services as a way to encourage antibiotic stewardship while safeguarding against any animal health risks associated with antibiotic use reduction. Farm-level trials and qualitative studies should be used to better examine the relationship between nontherapeutic antibiotic use and animal disease in this context, and to better understand how public veterinary services help to improve antibiotic stewardship and animal health outcomes, potentially supporting future interventions in which antibiotic use reduction is paired with an expansion of public veterinary services. These findings also raise questions about the incentives faced by non-public animal health service providers as they relate to antibiotic stewardship.

As always, we must remember that smallholder farmers form part of a complex network of actors, all of whom must be involved when designing and implementing antibiotic stewardship policies.

## Data Availability

The original and cleaned datasets, as well as the code used in our analyses, can be found on our GitHub. URL: https://github.com/Trescovia/AMUSE_Burkina_Faso

## Funding

This work was funded as part of the JPIAMR consortium SEFASI with funding for EE coming from the UK MRC, and funding for MD and AK coming from SIDA grant number APH002001. The views expressed are those of the author(s) and not necessarily those of the institutions the authors are affiliated with nor the funders of this research

## Notes

### Competing Interest Statement

The authors have declared no competing interest.

### Funding Statement

This work was funded as part of the JPIAMR consortium SEFASI with funding for EE coming from the UK MRC (UK Medical Research Council, https://www.ukri.org/councils/mrc/, grant code JPIAMR2021-182) and funding for MD and AK coming from SIDA (Swedish International Development Cooperation Agency, https://www.sida.se/en, grant code APH002001). The funders had no role in study design, data collection and analysis, decision to publish, or preparation of the manuscript.

### Author Declarations

The study was approved by the ethical committee of the Ministry of Health, Burkina Faso, with reference number 2020-9-186. Informed consent was obtained from each participant before they were interviewed. Consequently, all participants gave their consent to participate in the study.

